# Long-term sequelae of Pneumococcal Sepsis in children: a systematic review

**DOI:** 10.1101/2021.03.15.21253639

**Authors:** Adeteju Ogunbameru, Rafael Neves Miranda, Joanna Bielecki, Beate Sander

**Author notes:** **Corresponding author details:** Adeteju Ogunbameru, ^1^Institute of Health Policy, Management and Evaluation, University of Toronto, 155 College Street, Suite 425, Toronto, ON M5T 3M6, Canada. **Contributors:** AO and BS designed the study. JB developed the search strategy. AO and RM extracted and analyzed the data. AO drafted the manuscript. All authors revised the manuscript critically and gave final approval of the version to be published. AO is the guarantor. **Competing interests:** “All authors have completed the ICMJE uniform disclosure form at www.icmje.org/coi_disclosure.pdf and have no conflict of interest to declare. **Data sharing:** Study data were derived from published primary literature and are publicly available to other researchers. **Transparency declaration:** AO affirms that the manuscript is an honest, accurate, and transparent account of the study being reported; that no important aspects of the study have been omitted; and that any discrepancies from the study as originally planned (and, if relevant, registered) have been explained.

## Abstract

**Background:** Long-term sequelae associated with pneumococcal sepsis (PS) in pediatric patients in existing literature is currently unclear.

**Aim:** To review the evidence on sequelae and prognostic factors associated with PS among pediatric patients.

**Method:** We conducted a systematic review following the Preferred Reporting Items for Systematic Reviews and Meta-Analyses reporting guidelines. We screened six databases from their inception to January 15, 2021. Study population were neonates, infants, children and adolescents less than 18 years old with suspected or confirmed PS disease. Outcomes included sequelae types, prognostic factors, pooled death estimate and length of hospital stay (LOS) for survivors and deceased patients. Quality of studies was assessed using Joanna Briggs Institute appraisal checklists.

**Results:** We screened 981 abstracts, and 24 full-text articles for final review. Septic shock was the most prevalent physical sequelae reported (13%, n=1492 patients). No functional, cognitive or neurological sequelae were reported in included studies. Meta-analysis of pooled mortality estimate was 14.6% (95%CI: 9.9 −19.4%). Prognostic factors associated with increased risk of PS sequelae and death included pediatric risk of mortality score ≥ 10 and co-infection with meningitis. LOS for survivors and non-survivors ranged between 5-30 days and 1-30 days. Nine included studies met at least 50% of the quality assessment criteria.

**Conclusion:** Physical sequelae and death are the PS sequelae types currently identified in existing literature. Lack of information about other possible sequelae types suggests the long-term consequences of PS disease maybe underreported, especially in resource-limited settings. Future studies should consider exploring reasons for the existing of this knowledge gap.

## INTRODUCTION

Pneumococcus is one of the leading causes of child morbidity and mortality worldwide, accounting for approximately 335,000 (240,000–460,000) deaths in children aged less than 5 years in 2015.^1^ Infections of *S. pneumoniae* can lead to invasive syndromes such as meningitis, peritonitis and sepsis—and non-invasive syndromes which include acute otitis media and pneumonia. ^2^ Complications from pneumococcal sepsis (PS) can be severe and include mental retardation, hearing loss, multiorgan failure, neurological disorders, and death.^3^ Sepsis-related mortality, a known complication of acute infection illness has been vastly reported in literature ^4-6^. In a recent study,10% of childhood deaths under the age of 5 years in high-income countries are attributable to infections,^7^ however, other long-term physical, cognitive or psychological, and functional sequelae associated with pneumococcus sepsis are not well known.

An understanding of the sequelae and prognostic factors can inform clinical care and policy decision-making, including prevention strategies.^8^ To our knowledge, there has been no previously systematic review on the long-term sequelae of pneumococcal sepsis in children. Our aim is to conduct a systematic review to estimate the incidence of PS sequelae, prognostic factors and length of hospitalization associated with pneumococcal sepsis in pediatric patients.

## METHODS

Our systematic review protocol was registered in the International Prospective Register of Systematic Reviews (PROSPERO) database on March 23, 2020 (CRD 42020176275). The study reporting follows the Preferred Reporting Items in Systematic Reviews and Meta-Analyses (PRISMA) guideline. ^9^

### Search Strategy

All eligible studies were identified through a systematic comprehensive search of the Ovid MEDLINE(R) E-pub Ahead of Print, In-Process & Other Non-Indexed Citations, Ovid EMBASE, Ebsco CINAHL Plus, Psych Info and Cochrane Library databases for the period from inception of database to January 15, 2021. The search strategy was developed by an information specialist and it included Medical Subject Headings (MESH) terms and text words in the following concept areas: *Streptococcus pneumoniae*, sepsis, sequelae, prognostic factors and children. We developed the search strategy first in MEDLINE and subsequently translated it into other databases syntax (see Supplemental Digital Content 2 for MEDLINE search strategy). Retrieved studies from the five databases were imported into Endnote for de-duplication and transferred into Covidence for abstract and full text screening. Our inclusion criteria included cohort and randomized control trials designs, case-control, cross-sectional, case series with more than 10 patients. We excluded grey literature and opinion pieces (2) non-English articles with no English translation print at the University of Toronto library, and (3) animal studies. In addition, we manually searched through the reference lists of full text articles and other relevant studies like systematic reviews for additional studies. Data extraction was performed using Microsoft Excel.

### Selection criteria

Our systematic review included studies of patients under 18 years of age and had suspected or confirmed pneumococcal sepsis. Studies with mixed age population where the results of the children group were demarcated from the adult population were included. We excluded studies focused on the adult population. The exposure of interest was pneumococcal sepsis. The comparator of interest was patients without the syndrome of interest; however, the lack of a comparator in a study was not an exclusion criterion. For consistency, we defined pneumococcal sepsis as systemic inflammatory response syndrome (SIRS) as a result of suspected or proven pneumococcal infection.^10^ SIRS is defined by the International Pediatric Sepsis Consensus Task-Force as the presence of at least two of four conditions of which at least one must be abnormal temperature or abnormal leukocyte count. The four conditions are core temperature > 38.5°C and <36° C, tachycardia-defined as a mean heart rate greater than two standard deviations (SD) above normal for age in the absence of external stimulus, chronic drugs, or painful stimuli or unexplained persistent elevation over a 0.5hr to 4hr time period, mean respiratory rate greater than two SD above normal for age or mechanical ventilation for an acute process not related to underlying neuromuscular disease or general anesthesia, and white blood cell elevated or greater than 10% immature neutrophils.^10^ Long-term PS sequelae were defined the persistent presence of PS-related symptoms, complications and disabilities following hospital discharge or PS-related complications developed during hospitalization. We included studies that reported at least one of the following sequelae types: physical, neurological, cognitive or psychosocial, mortality and functional.^8^ We also included studies that reported prognostic factors and length of hospital stays.

### Study selection and Data extraction

All stages of the systematic review (i.e., title and abstract screening, full-text screening, data extraction and quality assessment) were conducted independently by two reviewers (AO and RM). Disagreements were resolved by consensus. We recorded reasons for exclusion of articles at full text screening. We extracted data on study and patient characteristics such as: study year, country of study, study design, sample size, sample of patients with PS, duration of follow-up, funding sources, previous antibiotic treatment, vaccination history, study age population and study outcomes examined.

### Quality of studies assessment

Two reviewers assessed the quality of studies independently using the Joanna Briggs Institute (JBI) appraisal checklists for cohort and randomized controlled trials.^11^ Studies were deemed of good quality by the research team if at least 50% of the quality assessment criteria were met and high quality if at least 80% of the quality assessment criteria were met for the given study design. Risk of bias assessment is presented in graphical format.

### Data synthesis and Meta-analysis

We described study and patient characteristics as well as sequelae types. We report the proportion of sequelae in the PS-diagnosed cohort of each study. Since mortality associated with PS was an outcome measured in several included studies, we conducted a meta-analysis to estimate the pooled proportion of deaths among subjects with PS using a random-effects model. ^12^ Studies were included for meta-analysis if death was recorded as a sequalae and if the sample size of subjects with PS was at least 10. Meta-analysis was conducted using the Metafor package in R statistical software. Summary of Q-value, degree of freedom (df), p-value and I^2^ values were used to determine the level of heterogeneity across included studies. For prognostic factors, we extracted odds ratio (with 95% confidence interval) for identified risk factors. For length of hospitalization, we extracted means and standard deviations (in days) associated with LOS for all patients and LOS from PS diagnosis until death for those who died. Additionally, results of study outcomes were stratified by the World Bank country classification (i.e., high income countries or low-and-middle income countries (LMIC)).

## RESULTS

Our search resulted in 1,915 studies from the five databases, of which 934 duplicates were removed. Through title and abstract screening, 870 studies were excluded because they did not meet inclusion criteria. The full text of 111 abstracts was assessed for further evaluation, of which 87 articles were excluded; the most common reasons for exclusions were: no specific data on Strep. pneumoniae induced sepsis (n= 21), case reports and case series with less than 10 patients (n=22), and no sequalae, prognostic factor or length of hospital data recorded (n= 15). We were unable to assess the full text of four studies, these studies were therefore excluded from our final analyses. The final review included 24 studies for the narrative synthesis of which 14 studies were included in the meta-analysis.^4,13-35^ The PRISMA flow chart is presented in Figure1. Studies and Patients characteristics

**Figure 1.**
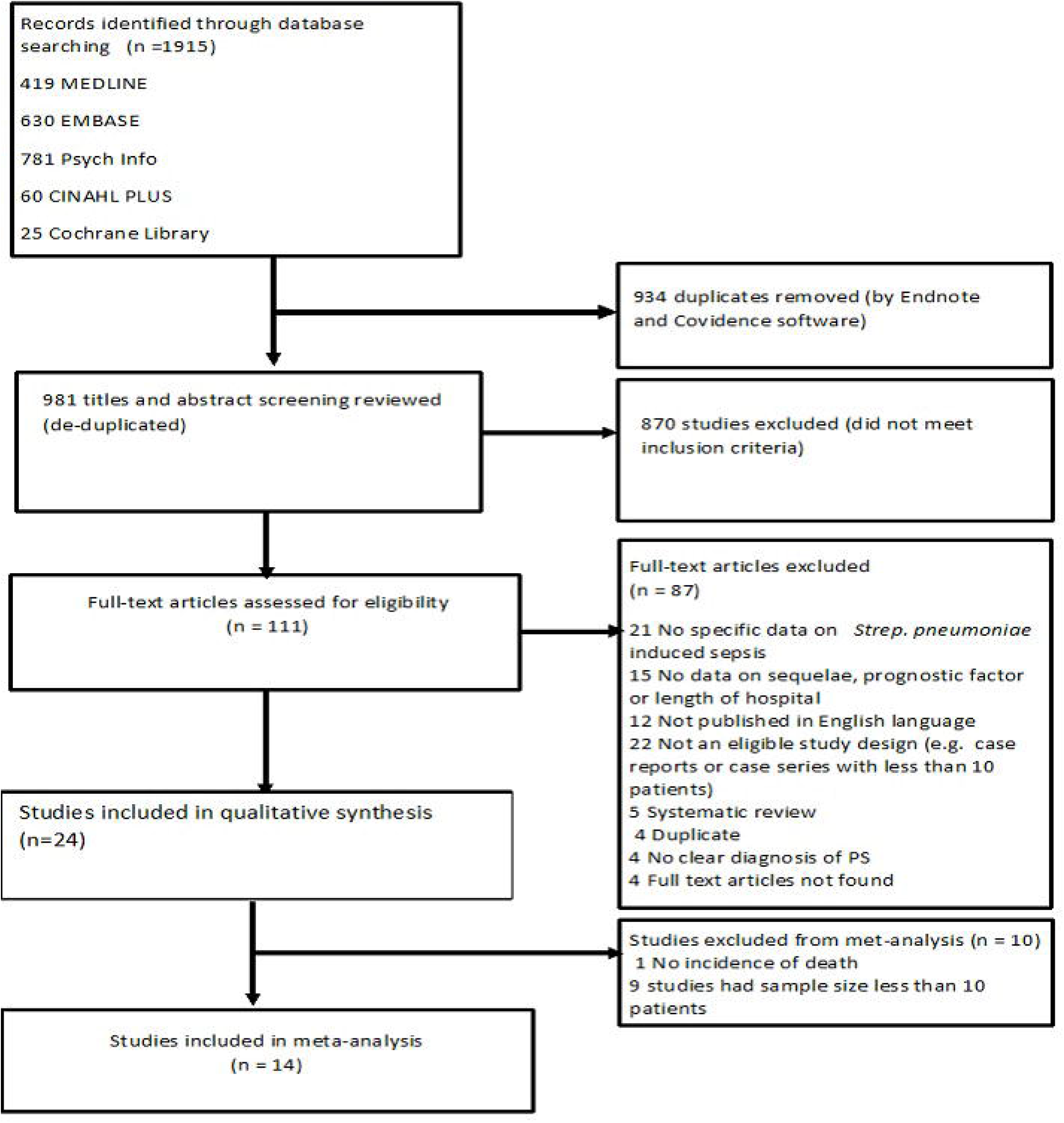
Preferred Reporting Items for Systematic Reviews and Meta-Analyses flow chart and study selection.

Studies were conducted between 1979 and 2020. Of the 24 studies, 17 were conducted in high income countries and 7 studies in LMIC. Countries where studies were conducted included: USA (6 studies), ^13-18^ China (3 studies), ^19-21^ Saudi Arabia (2 studies) ^22,23^ and Spain (2 studies), ^24,25^ and one study each in Kenya,^26^ Pakistan, ^27^ Chile,^28^ Taiwan,^29^ Germany,^30^ Italy,^31^ Switzerland, ^32^ Uruguay,^33^ Costa Rica ^34^ and Peru.^35^ One study was conducted in seven different European countries.^4^ Twenty-three studies used cohort designs (retrospective = 11, prospective = 11, retrospective and prospective =1) and one study used the randomized controlled trial design.^14^ Diagnosis of PS was confirmed by positive blood culture in all studies. Study sample size of patients diagnosed with PS ranged from 6 to 117. The age of the study population ranged from 0 to 17 years. The proportion of male study participants ranged from 53% to 71%. Duration of follow up ranged from 24 hours to 6 years for prospective and 5 to 16 years for retrospective studies. A detailed description of study and patient characteristics is described in Table 1.

**Table 1:** Study and Patient Characteristics

| **Author, Year** | **Country** | **Study Design** | **Age group** | **Mean age, ±SD** | **Sex, Male (%)** | **Study population size (% of PS patients)** | **Comorbidities reported** |
| --- | --- | --- | --- | --- | --- | --- | --- |
| Al Ayed, 2011 | Saudi Arabia | Cohort (Retrospective) | 0 – 12 years | NR | NR | 41 (0.05) | Chronic lung disease, Congenital heart disease, Renal diseases + Nephrotic syndrome, Splenic dysfunction, Sickle cell disease and Immuno-deficiency state |
| Asner, 2019 | Switzerland | Cohort (Prospective) | 0 – 17 years | NR | 57.2 | 117 (100) | Pneumonia (51%), Meningitis (25%), and Bloodstream infection (16%) |
| Assandri, 2015 | Uruguay | Cohort (Retrospective) | 0 - 28 days | NR | NR | 25 (84.0) | NR |
| Baron, 1989 | USA | Cohort (Prospective and Retrospective) | 3 - 24 months | NR | 52.6 | 189 (10.0) | NR |
| Bhutta, 1997 | Pakistan | Cohort (Prospective) | 0 - 30 days | Survivors: 6.5 (6.6), Non-survivors: 7.9 (8.6) | 66.8 | 292 (1.0) | NR |
| Boeddha, 2018 | 7 European Countries | Cohort (Prospective) | Less than 1 month -18 years | NR | 55 | 795 (9.8) | Meningitis/encephalitis (66%)<br>Pneumonia (27%) |
| Brent, 2006 | Kenya | Cohort (Prospective) | 0 – 5 years | 3 years | NR | 1093 (0.0009) | Human Immunodeficiency virus |
| Cai, 2018 | China | Cohort (Prospective) | 0 – 14 years | NR | 70.7 | 123 (66.6) | NR |
| Chiu, 2017 | Taiwan | Cohort (Retrospective) | 0 -17 years | NR | 60 | 50 (56) | NR |
| Gangoiti, 2018 | Spain | Cohort (Prospective) | 0-14 years | NR | 56.5 | 223 (11.8) | NR |
| Gaston, 1986 | USA | Randomized controlled trial | 3 -36 months | NR | 50 | 215 (7.4) | Waterhouse Frederickson syndrome, Pneumonia, Meningitis, Otitis media and Disseminated Intravascular coagulation |
| Hartman, 2013 | USA | Cohort (Retrospective) | 0 – 19 years | Year 1995: 4.8 (6.4); Year 2000:4.4 (6.5); Year 2005: 3.8 (6.3) | Year 1995: 54.2; Year 2000: 54.8; Year 2005: 55.8 | 14433 (2.3) | NR |
| Hoffman, 2003 | USA | Cohort (Prospective) | less than 30 days | 18.1 days (8.2) | 66 | 29 (27.6) | Respiratory syncytial virus, HIV, small bowel obstruction/hernia repair, bronchiolitis, and Maternal Group B Streptococcus colonization |
| Hongeng, 1997 | USA | Cohort (Retrospective) | 0-5 years | 21.5 months | 62.5 | 8 (100) | Sickle cell Disease |
| Lagos, 2008 | Chile | Cohort (Prospective) | 0 - 14 years | NA | 45 | 2369 (4.9) | NR |
| Li, 2019 | China | Cohort (Retrospective) | 0 -17 years | NR | 59.3 | 172 (100) | Immunosuppression (13.9%), Hematologic malignancy (0.05%), Congenital heart disease, (0.03%) |
| Li, 2019b | China | Cohort (Retrospective) | 0 – 17 years | NR | 58.1 | 136 (27.9) | Immunosuppression, Congenital heart disease, Hematologic malignancy |
| Memish, 2010 | Saudi Arabia | Cohort (Retrospective) | 0 – 5 years | NR | 53.9 | 82 (76.8) | NR |
| Ochoa, 2010 | Peru | Cohort (Prospective) | 0 – 16 years | NR | 48.4 | 101 (7.9) | NR |
| Powars, 1981 | USA | Cohort (Prospective) | 0 – 6 years | NR | NR | 233 (14.5) | sickle cell anemia |
| Resti, 2009 | Italy | Cohort (Prospective) | 0 – 16 years | 4.6 years | 60 | 83 (8.4) | NR |
| Schnappauf 2014 | Germany | Cohort (Retrospective) | 0 – 18 years | NR | 62 | 55 (36.4) | Interleukin-1 receptor-associated kinase 4 (IRAK 4) deficiency, (0.1%), meningitis (0.4%) and pneumonia with effusion (0.15%) |
| Sole, 2018 | Spain | Cohort (Retrospective) | 0 – 18 years | NR | 66.6 | 21 (23.8) | NR |
| Ulloa-Gutierrez, 2003 | Costa Rica | Cohort (Retrospective) | 0-11.4 years | 35.7 months | 59.3 | 132 (22.7) | Bacteremia (46.7%) |

NR, Not reported; PS, Pneumococcal Sepsis; SD, standard deviation.

All 24 studies reported sequelae in frequencies and proportions. Nine studies reported physical sequelae, ^4,13,14,16,18,23,25,30,32,^ 16 studies reported death ^4,14,15,18-21,23,28-30,32-34,35^ and 19 studies reported discharge outcomes. ^4,13,14,16-25,28,229,31,32,34-35^ The type and proportion of physical sequelae is presented as a graph. Prognostic factors were reported in 5 studies. ^20,21,24,27,32^ Quality Appraisal

Nine studies (37.5%) met at least 50% of the quality assessment criteria; 8 (33%) met at least 80% of their respective criteria. In particular, eight (72%) of the 11 prospective cohort studies and the only randomized controlled trial study met 80% of the quality assessment criteria. Retrospective cohort studies met less than 50% criteria, with the score ranging from 27% to 45%. (Figure 2) Among cohort studies, the domain most commonly unmet was confounder identification.

**Figure 2.**
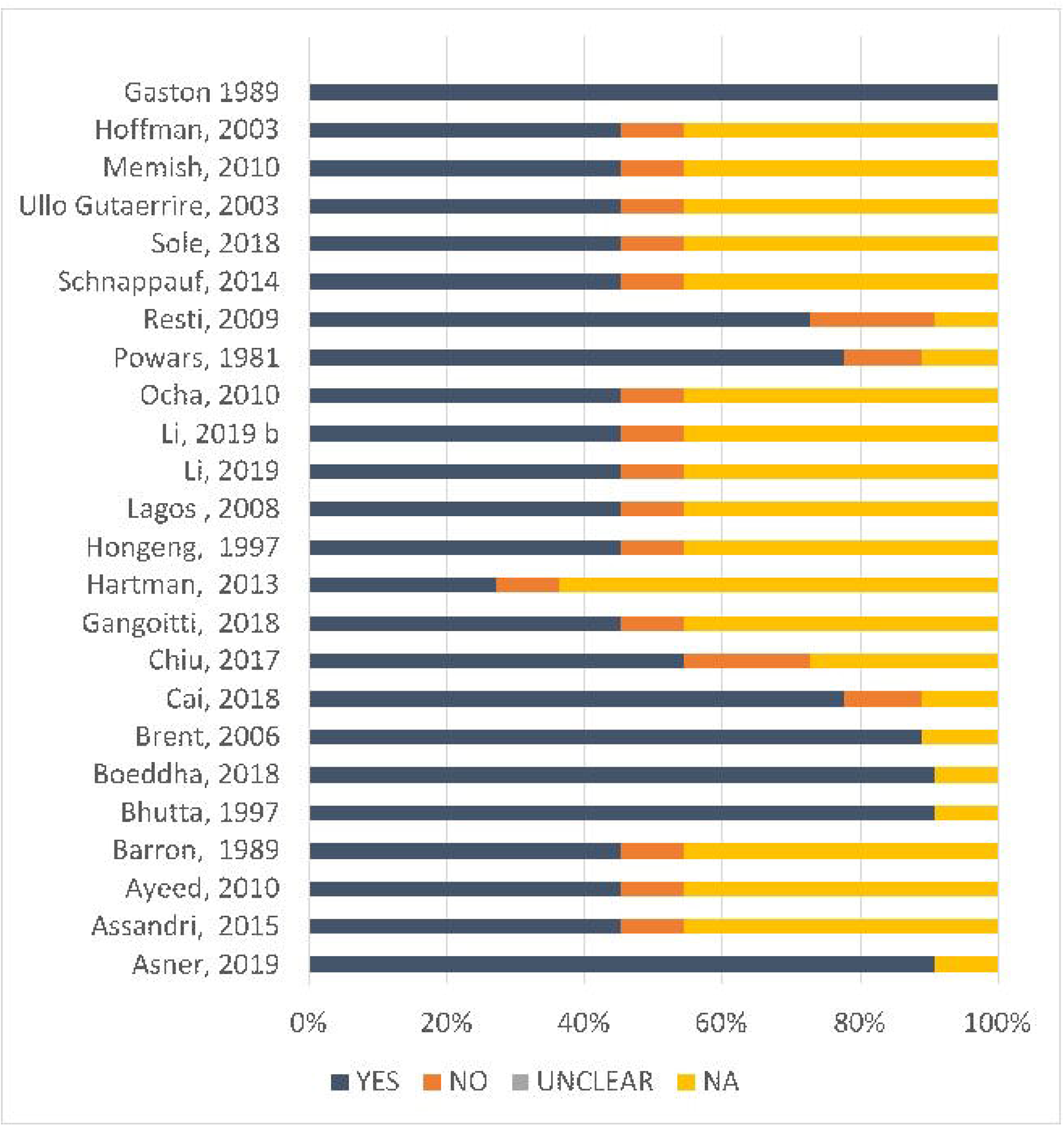
Quality appraisal results using JBI checklist for cohort and randomized controlled studies ^1^ ^1^The tool evaluates quality using five main domains which include study participants selection, exposure and outcome identification and measurement, confounders identification and statistical analysis. Possible scores for the different domains range from Oto 3 for ‘No’, ‘Unclear’, ‘Not applicable’ and ‘Yes’ responses respectively. Based on the study design, the percentage of positive responses (i.e., “Yes”) of each study was computed.

### Sequelae

In a quantitative analysis, 9 studies conducted in high income countries reported physical sequelae. No physical sequelae were reported in LMIC studies. We stratified by type of sequelae and summarized the percentage of each type of sequelae reported.(Figure 3) In studies from high income countries, septic shock, ^4,14,16,25,32^ severe sepsis associated with organ dysfunction (e.g., chronic renal failure, cardiovascular disease, end organ renal failure) ^4,32,32^ and persistent infection (such as periorbital cellulitis and meningitis) ^4,13,23^ were reported as common physical sequelae associated with PS infection with septic shock being the most prevalent physical sequelae, occurring in 194 of`1492 PS-infected patients (13%). Less common physical sequelae included amputation (3.4%) and splenic sequestration (1.0%).

**Figure 3:**
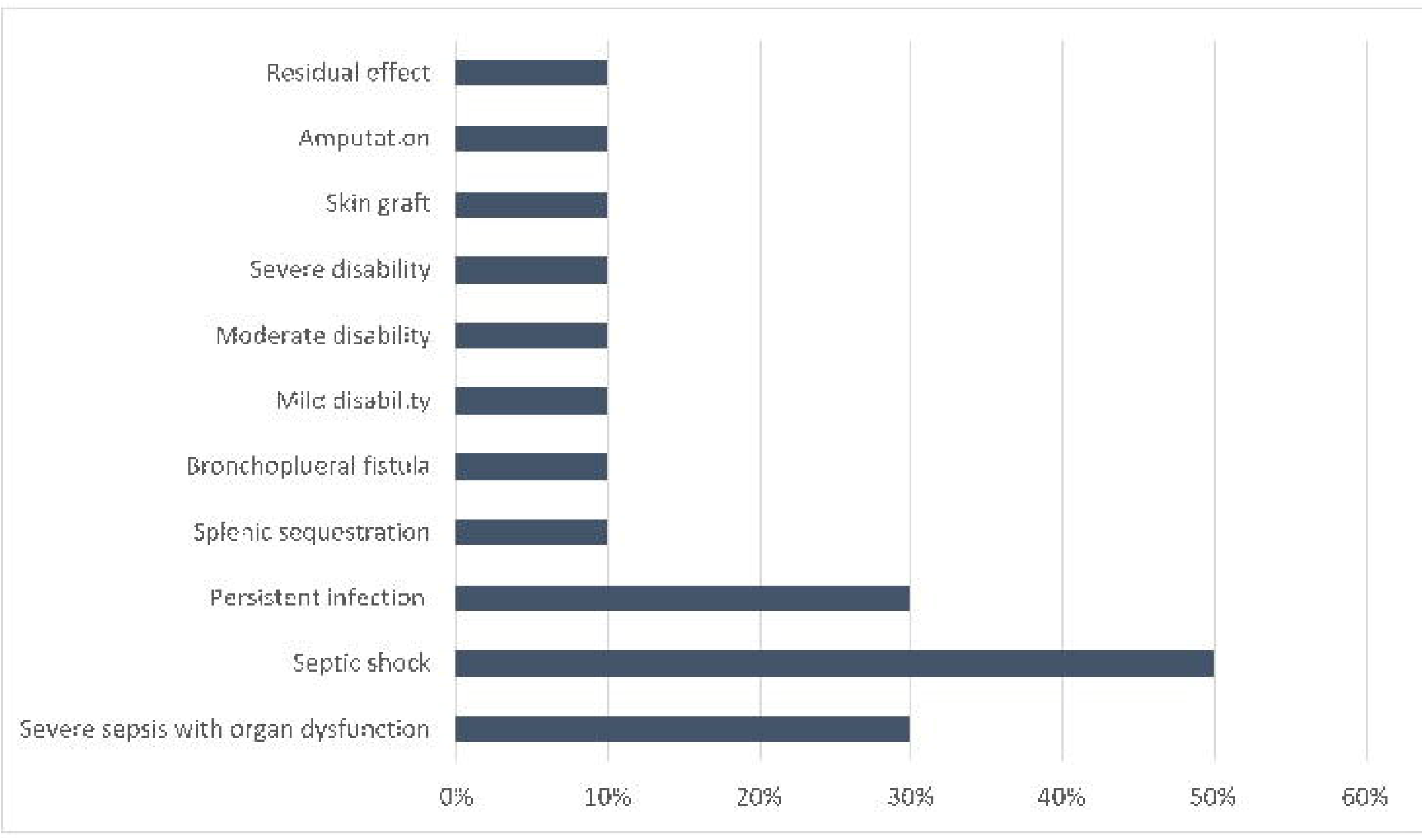
Types and Proportions of physical sequalae

For mortality, results of the meta-analysis showed a high level of heterogeneity across included studies (n =14) (Figure 4). The test of heterogeneity revealed I^2^ value of 81.8% (p < 0.001), Q value of 72.13, and 13 degrees of freedom (df) (Q-df = 59.1). Given the high-level heterogeneity across studies, mortality results were further summarized using descriptive synthesis. Meta-analysis of pooled mortality estimate was 14.6% (95%CI 9.9 −19.4%). The overall mortality rate across studies ranged from 3.6%-50%. Prior to 2010 (i.e., before development of the pneumococcal vaccine), the reported mortality ranged from 6.7% to 23% in high-income country settings ^14,18,33^ but decreased after the introduction of the vaccine (3.6% - 20%). ^4,15,23,29,30,33^ In LMIC settings, one study reported a mortality of 28.8% ^28^ prior to 2010. Post 2010, the reported mortality ranged from 3.65% to 50%. ^19-21,32^ Time to death after PS diagnosis ranged from less than 9 hours to 30 days across 19 studies.

**Figure 4.**
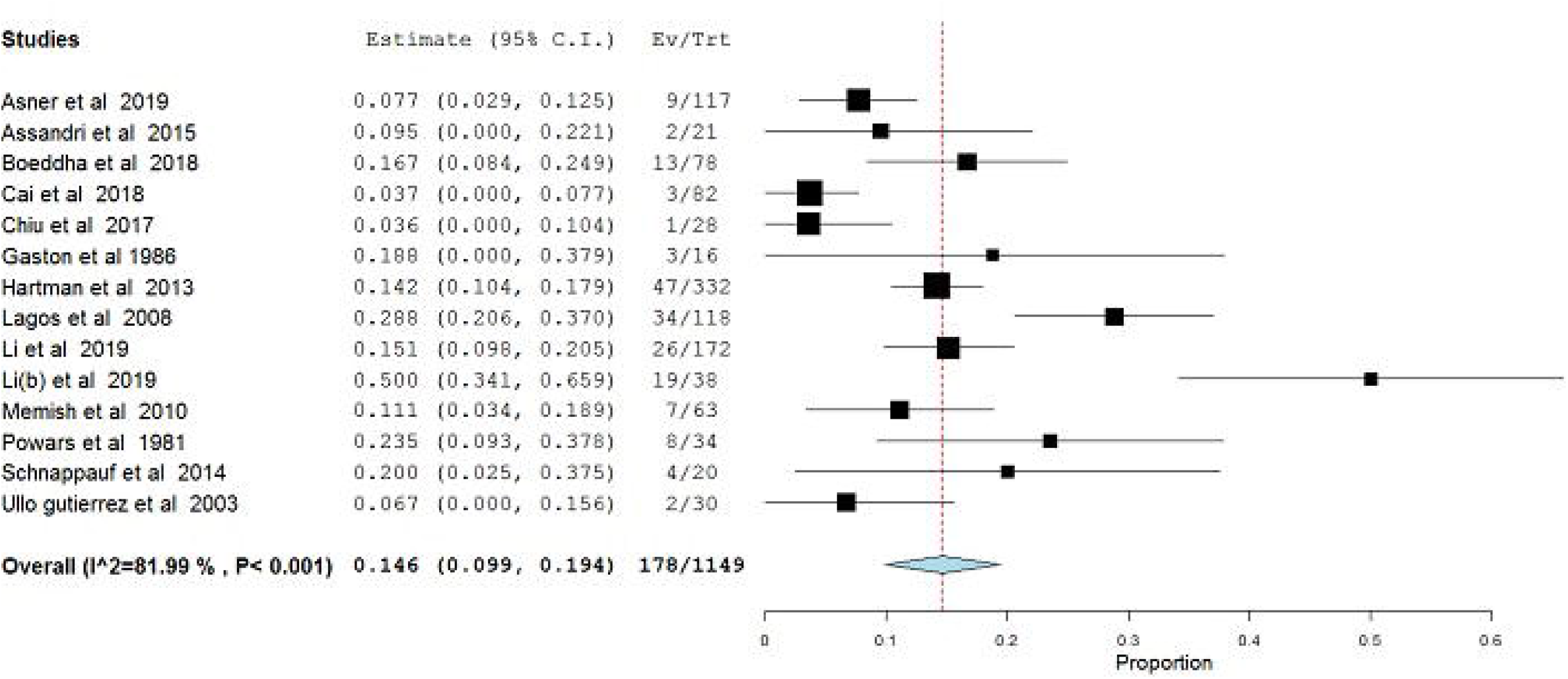
Forest plot showing pooled mortality estimate

### Prognostic factors

Five studies reported prognostic factors for PS associated sequelae and in-hospital mortality using unadjusted and adjusted odds ratio estimates. ^20,21,24,27,32^ Two of the studies were from high income countries (Switzerland and Spain), ^24,32^ and remaining three from low- and middle-income countries (China and Pakistan). ^20,21,27^ All five studies provided adjusted OR estimates to control for confounders which included age, sex, presence of underlying diseases and infections, vaccine failure and child weight at birth. From multivariate analysis results of four studies, risk factors associated with an increased risk of PS physical sequelae included co-infection with meningitis ^32^ and duration of fever < 24 hours.^24^ Risk factors associated with PS death included hypotensive shock ^27^ and living in impoverished settings ^24^. In general, a pediatric risk of mortality score ≥ 10 had the highest adjusted OR of 51.08 [95% 8.19-318.54] for a fatal outcome.^24^ PS-diagnosed patients co-infected with meningitis were seven times more likely to experience septic shock [OR: 6.8; 95%CI 2.4 −19.3] compared to patients without meningitis.^32^ Length of hospitalization

Eleven studies reported on LOS. Five studies were conducted in LMIC ^19,21^,^22,27^ and six in high income countries. ^4,10,17,23,31-33^ Overall LOS for survivors was between 5 to 30 days. LOS until death, reported in high income studies, ranged from less than 1 day to 30 days. ^24,27,32,35^ In LMIC settings, the mean LOS until death (in days) reported was between 5 to 7 days.^27,35^ One study conducted in high income countries reported on risk factors that prolong overall LOS, which included infection with *Streptococcus pneumoniae* serotype 3 and presence of meningitis as co-infection. ^32^

### Recovery outcomes

Nineteen studies reported discharge outcomes with no clear specification on recovery status (i.e., full or partial).^4,10,17,20-24,26-34,36^ Among the 19 studies, 80% of patients (n= 905) recovered after treatment. In two studies that reported physical sequelae, about 8% of patients (n=63) had “residual effect” in one study ^23^ and 10% (n= 30) experienced “residual effect” in another after hospital discharge.^34^

## DISCUSSION

Our systematic review summarized evidence on pneumococcal sepsis-related long-term sequelae, prognostic factors and LOS in pediatric patients. In total, we included 24 studies published until January 15, 2020. Thirty-three percent of included studies were conducted after 2010, when the pneumococcal vaccine was developed, indicating that pneumococcal sepsis remains of high interest world-wide. Among these studies, half were conducted in low- and middle-income countries, which continue to have a high incidence of pneumococcal sepsis ^[36]^ given limited access to pneumococcal vaccine. ^37^ While previously published cohort studies on invasive pneumococcal disease have identified long-term sequelae associated with pneumococcal sepsis, no systematic review has synthesized the evidence on long-term sequelae of pneumococcal sepsis.

The overall quality of the studies was heterogeneous. Only eight studies met 80% of the JBI checklist domains - seven cohort studies and one randomized controlled trial study. More than half of the studies failed to identify confounding variables.

Septic shock was the most common physical sequelae. Septic shock has previously been identified as a common complication of pneumococcal sepsis in children.^32^ Mortality due to pneumococcal sepsis has been reduced globally post-approval of the pneumococcal vaccine. Overall, mortality is still higher in LMIC compared to high income countries. The higher mortality rate has been attributed to poor critical care services, ^38^ and limited health resources in these settings. ^39^ The high level of heterogeneity in our meta-analysis analysis result can be attributed to various reasons such as difference, study designs settings and patient characteristics across included studies.

Sequelae occurred mostly among children with underlying medical conditions, and patients with a Pediatric Risk of Mortality score greater than 10 were at highest risk. Presence of co-infection (specifically meningitis) and system dysfunction disease (neurological, respiratory) were associated with early mortality. One risk factor noteworthy to increase septic shock and mortality is the availability of mechanical ventilation in pediatric intensive care units.^21^ Ventilators, a breathing-support equipment, is scarcely available in many hospitals in LMIC settings. Thus, to reduce physical and death sequelae in LMIC, it is imperative to develop policies that support hospital critical care units with advanced medical equipment like ventilators, and fund vaccination programs.

Our review has some limitations. In an attempt to generate reliable evidence, we excluded some study designs such as case reports and case series with less than 10 study participants, which inadvertently may have excluded some sequelae. Studies included in our meta-analysis had varying follow-up durations which could potentially confound our findings. Our findings could also be subject to language bias because we only studies published in English language in peer-reviewed journals. Because we did not search the grey literature, our results might be subject to publication bias towards a positive result and we might have excluded some studies reporting on PS sequelae outcomes.

To our knowledge, this is the first systematic review that estimate the incidence of PS sequelae in pediatric patients. Our finding is strengthened by our inclusion of multiple databases during review to generate a detailed and comprehensive evidence. We consulted an information expert to ensure search concepts was robust and approximate. We placed no restriction on study setting or publication dates in our search strategy.

With the current global interest to improve sepsis management,^40^ the findings of the review can help inform decision-makers at national and international levels on how to channel economic and clinical resources to improve health outcomes in PS patients.

## CONCLUSION

Our review provides a comprehensive assessment of the long-term sequelae and risk factors for pneumococcal sepsis infection. Physical sequelae and death are the PS sequelae types currently reported in existing literature. This suggest that there’s possibility of underreporting of other sequelae types and future research could explore reasons for this existing knowledge gap. The prognostic factors associated with these sequelae included risk of mortality score ≥ 10, presence of co-infection during illness and living in impoverished settings.

## Data Availability

All data were obtained from primary literature and are publicly available to other researchers.

## Acknowledgements

The authors thank Dr. Lusine Abrahamyan, Assistant Professor and scientist, Toronto Health Economics and Technology Assessment (THETA) Collaborative, University Health Network for her technical assistance during the study conceptualization.

## List of Abbreviations

LOS: Length of Hospital stay
LMIC: Low-and-middle-income countries
PS: Pneumococcal sepsis

