## Appendix 1 for "Long-term sequelae of Pneumococcal Sepsis in children: a systematic review"

**APPENDIX A: PRISM REPORTING CHECKLIST**

| **Section/topic** | **#** | **Checklist item** | **Reported on page #** |
| --- | --- | --- | --- |
| **TITLE** | | |  |
| Title | 1 | Identify the report as a systematic review, meta-analysis, or both. | 1 |
| **ABSTRACT** | | |  |
| Structured summary | 2 | Provide a structured summary including, as applicable: background; objectives; data sources; study eligibility criteria, participants, and interventions; study appraisal and synthesis methods; results; limitations; conclusions and implications of key findings; systematic review registration number. | 2 |
| **INTRODUCTION** | | |  |
| Rationale | 3 | Describe the rationale for the review in the context of what is already known. | 3 |
| Objectives | 4 | Provide an explicit statement of questions being addressed with reference to participants, interventions, comparisons, outcomes, and study design (PICOS). | 4 |
| **METHODS** | | |  |
| Protocol and registration | 5 | Indicate if a review protocol exists, if and where it can be accessed (e.g., Web address), and, if available, provide registration information including registration number. | 4 |
| Eligibility criteria | 6 | Specify study characteristics (e.g., PICOS, length of follow-up) and report characteristics (e.g., years considered, language, publication status) used as criteria for eligibility, giving rationale. | 4 - 5 |
| Information sources | 7 | Describe all information sources (e.g., databases with dates of coverage, contact with study authors to identify additional studies) in the search and date last searched. | 6 |
| Search | 8 | Present full electronic search strategy for at least one database, including any limits used, such that it could be repeated. | 6 |
| Study selection | 9 | State the process for selecting studies (i.e., screening, eligibility, included in systematic review, and, if applicable, included in the meta-analysis). | 6 |
| Data collection process | 10 | Describe method of data extraction from reports (e.g., piloted forms, independently, in duplicate) and any processes for obtaining and confirming data from investigators. | 6 |
| Data items | 11 | List and define all variables for which data were sought (e.g., PICOS, funding sources) and any assumptions and simplifications made. | 6-7 |
| Risk of bias in individual studies | 12 | Describe methods used for assessing risk of bias of individual studies (including specification of whether this was done at the study or outcome level), and how this information is to be used in any data synthesis. | 7 |
| Summary measures | 13 | State the principal summary measures (e.g., risk ratio, difference in means). | 7 |
| Synthesis of results | 14 | Describe the methods of handling data and combining results of studies, if done, including measures of consistency (e.g., I^2^) for each meta-analysis. | 7 |

| **Section/topic** | **#** | **Checklist item** | **Reported on page #** |
| --- | --- | --- | --- |
| Risk of bias across studies | 15 | Specify any assessment of risk of bias that may affect the cumulative evidence (e.g., publication bias, selective reporting within studies). | 7 |
| Additional analyses | 16 | Describe methods of additional analyses (e.g., sensitivity or subgroup analyses, meta-regression), if done, indicating which were pre-specified. | 7 |
| **RESULTS** | | |  |
| Study selection | 17 | Give numbers of studies screened, assessed for eligibility, and included in the review, with reasons for exclusions at each stage, ideally with a flow diagram. | 8; 10 |
| Study characteristics | 18 | For each study, present characteristics for which data were extracted (e.g., study size, PICOS, follow-up period) and provide the citations. | 8-9; 11-13 |
| Risk of bias within studies | 19 | Present data on risk of bias of each study and, if available, any outcome level assessment (see item 12). | 14-15; 19 |
| Results of individual studies | 20 | For all outcomes considered (benefits or harms), present, for each study: (a) simple summary data for each intervention group (b) effect estimates and confidence intervals, ideally with a forest plot. | 15-17 |
| Synthesis of results | 21 | Present results of each meta-analysis done, including confidence intervals and measures of consistency. | 17, Appendix D |
| Risk of bias across studies | 22 | Present results of any assessment of risk of bias across studies (see Item 15). | 14 |
| Additional analysis | 23 | Give results of additional analyses, if done (e.g., sensitivity or subgroup analyses, meta-regression [see Item 16]). | NA |
| **DISCUSSION** | | |  |
| Summary of evidence | 24 | Summarize the main findings including the strength of evidence for each main outcome; consider their relevance to key groups (e.g., healthcare providers, users, and policy makers). | 19 - 20 |
| Limitations | 25 | Discuss limitations at study and outcome level (e.g., risk of bias), and at review-level (e.g., incomplete retrieval of identified research, reporting bias). | 20-21 |
| Conclusions | 26 | Provide a general interpretation of the results in the context of other evidence, and implications for future research. | 21 |
| **FUNDING** | | |  |
| Funding | 27 | Describe sources of funding for the systematic review and other support (e.g., supply of data); role of funders for the systematic review. | 21 |

**SEARCH STRATEGY IN MEDLINE**

**OvidMEDLINE (R) :**1946 to February 13, 2020
Search Strategy:

| **#** | **Searches** | **Results** |
| --- | --- | --- |
| 1 | sepsis/ or neonatal sepsis/ or shock, septic/ or exp bacteremia/ or (seps?s or (blood adj2 poisoning?) or py?emia? or pyohemia? or Septicemia? or Bacteremia? or endotoxemia? or ((Septic or Toxic or Endotoxic) adj3 Shock)).ti,ab,kf. | 168857 |
| 2 | Streptococcus pneumoniae/ or Pneumococcal Infections/ or ("Streptococcus pneumoniae" or "Strep. Pnemo" or "S. pneumoniae" or "Strep. Pneumoniae" or "Diplococcus pneumoniae" or Pneumococc$ or (Pneumococc$ adj3 (Disease? or Infection?))).ti,ab,kf. | 42397 |
| 3 | (((coexistent or co-existing or associated or concomitant) adj4 (disease? or condition?)) or sequelae or sequels or complication?).ti,ab,kf. | 1057800 |
| 4 | prognosis.sh. or [diagnosed.tw](http://diagnosed.tw/). or cohort:.mp. or predictor:.tw. or [death.tw](http://death.tw/). or exp models, statistical/ | 2364207 |
| 5 | 3 or 4 | 3206987 |
| 6 | exp *adolescence/ or exp *adolescent/ or exp *child/ or exp *childhood disease/ or exp *infant disease/ or (adolescen* or babies or baby or boy? or boyfriend or boyhood or girlfriend or girlhood or child or child* or child*3 or children* or girl? or infan* or juvenil* or juvenile* or kid? or minors or minors* or neonat* or neo-nat* or newborn* or new-born* or paediatric* or peadiatric* or pediatric* or perinat* or preschool* or puber* or pubescen* or school* or teen* or toddler? or underage? or under-age? or youth*).ti,kw. | 1464877 |
| 7 | 1 and 2 and 5 and 6 | 355 |
| 8 | limit 7 to (address or autobiography or bibliography or biography or comment or congress or consensus development conference or consensus development conference, nih or dictionary or directory or editorial or "expression of concern" or festschrift or government document or guideline or historical article or interactive tutorial or lecture or legal case or legislation or letter or news or newspaper article or overall or patient education handout or periodical index or personal narrative or portrait or practice guideline or published erratum or retracted publication or "retraction of publication" or video-audio media or webcasts) | 5 |
| 9 | 7 not 8 | 350 |
| 10 | animal/ not (animal/ and human/) | 4640722 |
| 11 | 9 not 10 | 348 |

**APPENDIX C: Assessment of Quality using JBI**

**For cohort studies**

| **Study** | **Q1** | **Q2** | **Q3** | **Q4** | **Q5** | **Q6** | **Q7** | **Q8** | **Q9** | **Q10** | **Q11** |
| --- | --- | --- | --- | --- | --- | --- | --- | --- | --- | --- | --- |
| **Ayeed, 2010** | NA | NA | Y | NA | NA | N | Y | Y | Y | NA | Y |
| Asner, 2019 | Y | Y | Y | Y | Y | Y | Y | Y | Y | NA | Y |
| Assandri, 2015 | NA | NA | Y | NA | NA | N | Y | Y | Y | NA | Y |
| Barron, 1989 | NA | NA | Y | NA | NA | N | Y | Y | Y | NA | Y |
| Bhutta, 1997 | Y | Y | Y | Y | Y | Y | Y | Y | Y | NA | Y |
| Boeddha, 2018 | Y | Y | Y | Y | Y | Y | Y | Y | Y | NA | Y |
| Brent, 2006 | Y | Y | Y | U | U | Y | Y | Y | Y | NA | Y |
| Cai, 2018 | Y | Y | Y | U | U | Y | Y | Y | Y | NA | N |
| Chiu, 2017 | NA | NA | Y | Y | N | N | Y | Y | Y | NA | Y |
| Gangoitti, 2018 | NA | NA | Y | NA | NA | N | Y | Y | Y | NA | Y |
| Hartman, 2013 | NA | NA | Y | NA | NA | N | Y | NA | NA | NA | Y |
| Hongeng, 1997 | NA | NA | Y | NA | NA | N | Y | Y | Y | NA | Y |
| Lagos , 2008 | NA | NA | Y | NA | NA | N | Y | Y | Y | NA | Y |
| Li, 2019 | NA | NA | Y | NA | NA | N | Y | Y | Y | NA | Y |
| Li, 2019 b | NA | NA | Y | NA | NA | N | Y | Y | Y | NA | Y |
| Ocha, 2010 | NA | NA | Y | NA | NA | N | Y | Y | Y | NA | Y |
| Powars, 1981 | Y | Y | N | U | U | Y | Y | Y | Y | NA | Y |
| Resti, 2009 | Y | Y | Y | N | N | Y | Y | Y | Y | NA | Y |
| Schnappauf, 2014 | NA | NA | Y | NA | NA | N | Y | Y | Y | NA | Y |
| Sole, 2018 | NA | NA | Y | NA | NA | N | Y | Y | Y | NA | Y |
| Ullo Gutaerrire, 2003 | NA | NA | Y | NA | NA | N | Y | Y | Y | NA | Y |
| Hoffman, 2003 | NA | NA | Y | NA | NA | N | Y | Y | Y | NA | Y |
| Memish, 2010 | NA | NA | Y | NA | NA | N | Y | Y | Y | NA | Y |

*Y, Yes; N, No; U, Unclear; NA, Not applicable

Questions: 1. Were the two groups similar and recruited from the same population?; 2. Were the exposures measured similarly to assign people to both exposed and unexposed groups?; 3. Was the exposure measured in a valid and reliable way?; 4. Were confounding factors identified? 5. Were strategies to deal with confounding factors stated?; 6. Were the groups/participants free of the outcome at the start of the study (or at the moment of exposure)?; 7. Were the outcomes measured in a valid and reliable way?; 8. Was the follow up time reported and sufficient to be long enough for outcomes to occur?; 9. Was follow up complete, and if not, were the reasons to loss to follow up described and explored?; 10. Were strategies to address incomplete follow up utilized?; 11. Was appropriate statistical analysis used?

**For RCT study**

| **Study** | **Q1** | **Q2** | **Q3** | **Q4** | **Q5** | **Q6** | **Q7** | **Q8** | **Q9** | **Q10** | **Q11** | **Q12** |
| --- | --- | --- | --- | --- | --- | --- | --- | --- | --- | --- | --- | --- |
| Gaston 1986 | Y | Y | Y | Y | Y | Y | Y | Y | Y | Y | Y | Y |

*Y, Yes

Questions: 1. Was true randomization used for assignment of participants to treatment groups? 2 Was allocation to treatment groups concealed? 3.Were treatment groups similar at the baseline? 4. Were participants blind to treatment assignment? 5. Were outcomes assessors blind to treatment assignment?	6.Were treatment groups treated identically other than the intervention of interest? 7.Was follow up complete and if not, were differences between groups in terms of their follow up adequately described and analyzed?; 8. Were participants analyzed in the groups to which they were randomized?; 9.Were outcomes measured in the same way for treatment groups?; 10. Were outcomes measured in a reliable way?;	11. Was appropriate statistical analysis used?;12.Was the trial design appropriate, and any deviations from the standard RCT design (individual randomization, parallel groups) accounted for in the conduct and analysis of the trial?

**APPENDIX D: Results of Meta-analysis**

Binary Random-Effects Model

Metric: Proportion

Model Results

Estimate	Lower bound	Upper bound	Std. error	p-Value

0.146	0.099	0.194	0.024	< 0.001

Test of	Heterogeneity

tau^2 Q (df=13)	Het. p-Value	I^2

0.006	72.174	< 0.001	81.988

I^2^ = 81.9% indicates high level of heterogeneity (p < 0.001)

**FOREST PLOT OF META-ANALYSIS**

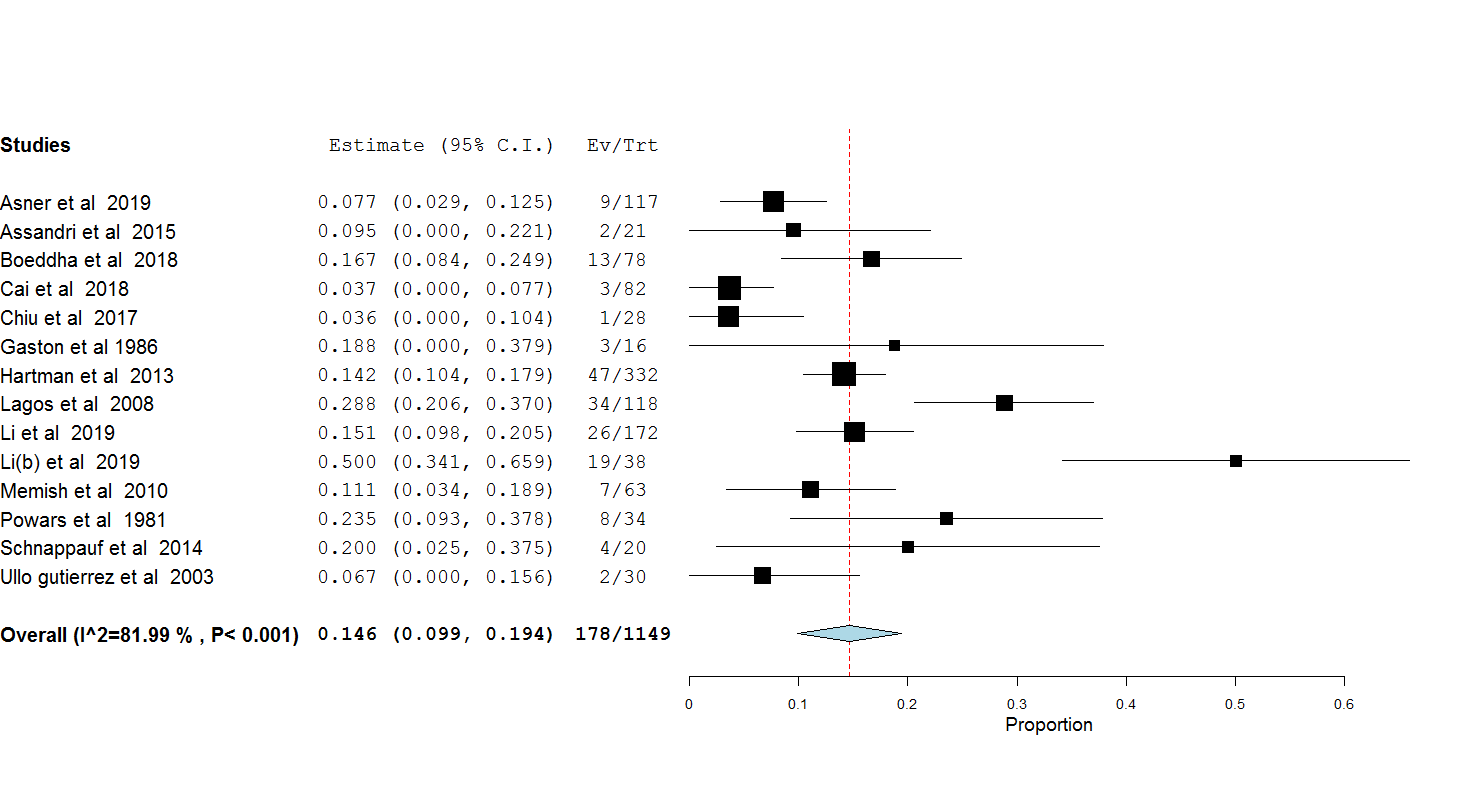

**FUNNEL PLOT OF META-ANALYSIS**

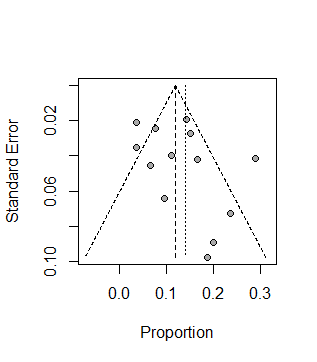

**NUMBER OF STUDIES WITH INFLUENCIAL OBSERVATIONS**

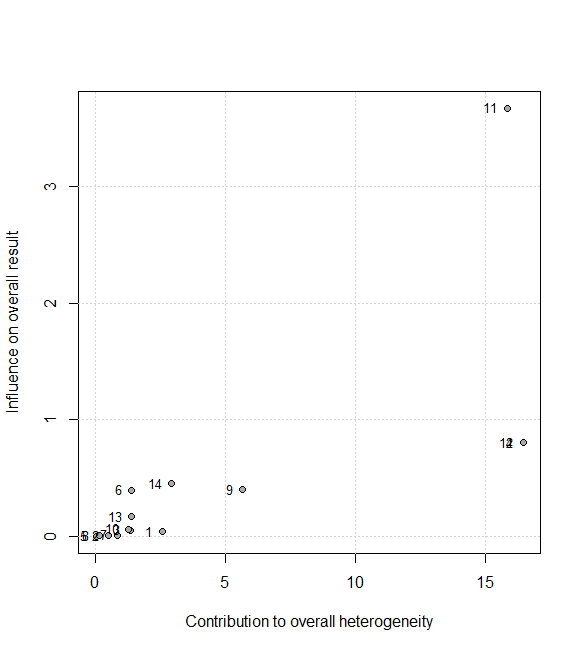

**11 studies had influential observations.**

**APPENDIX E: Prognostic factors analysis**

**Risk factors associated with in- hospital mortality**

| **Study** | **Risk factor** | **Adjusted OR** | **95% CI** | **Unadjusted OR** | **95% CI** |
| --- | --- | --- | --- | --- | --- |
| Bhutta 1997 [19] | Hypotensive shock | 3.59 | 1.34 – 9.62 | NR | NR |
|  | Renal failure | 11.23 | 4,43 – 29.05 | NR | NR |
|  | Respiratory failure | 8.43 | 3.30 – 24.54 | NR | NR |
| Li 2019 [29] | PRISM score ≥ 10 | 51.08 | 8.19 -318.54 | 38.81 | 12.97-116.16 |
|  | TTAT≥ 13.6 hrs. | 39.26 | 6.10 -252.60 | 21.35 | 7.33 – 62.18 |
|  | TTP ≤ 12 hrs. | 15.62 | 2.66 – 91.88 | 7.62 | 3.08 – 18.83 |
|  | Poverty stricken county residence | NR | NR | 7.74 | 3.15 – 19.04 |
|  | Empiric treatment with carbapenems | NR | NR | 8.67 | 1.82- 41.36 |
|  | Hematologic malignancy | NR | NR | 8.45 | 2.10 – 34.02 |
|  | Immunosuppression | NR | NR | 4.62 | 1.76 -12.18 |
|  | CNS infection | NR | NR | 4.24 | 1.78 – 10.12 |
| Li 2019 b [30] | Need for invasive mechanical ventilation | 20.52 | 3.50 – 120.47 | 24.61 | 8.03 – 75.38 |
|  | PRISM ≥ 10 | 4.31 | 1.04 – 11.79 | 34.61 | 10.72 – 111.76 |
|  | Early TTP | 18.91 | 3.36 – 106.59 | 8.21 | 3.08 – 21.89 |
|  | Intensive care unit admission | NR | NR | 12.25 | 4.44-33.8 |
|  | Meningitis |  |  | 4.30 | 1.69 -10.97 |
|  | Immunosuppression |  |  | 3.77 | 1.35 – 10.51 |

*PRISM – Pediatric mortality score, TTAT – time to appropriate therapy, TTP- time to positivity

**Risk factors associated with physical sequelae**

| Study | Risk factor | Adjusted OR | 95% CI | Unadjusted OR | 95% CI |
| --- | --- | --- | --- | --- | --- |
| Gangoiti 2018 | Duration of fever (< 24 hrs.) | 4.08 | 1.85 - 8.96 | NR | NR |
|  | Symptoms other than fever (neurological, respiratory) | 3.87 | 1.50 – 9.94 | NR | NR |
|  | Well-appearance (No) | 8.28 | 3.73 – 18.37 | NR | NR |
| Asner 2019 | Meningitis versus No meningitis | 6.8 | 2.4 -19.3 | 6.7 | 2.6 – 16.8 |
|  | Infants less than 12 months) | NR | NR | 2.6 | 1.04 - 6.5 |
| Li 2019 | PRISM III score ≥ 10 | 10.62 | 3.48 – 32.39 | 16.95 | 6.16 – 46.61 |
|  | TTP ≤ 12hrs | 4.02 | 1.65 – 9.75 | 5 | 2.38 – 10.52 |
|  | TTAT ≥ 13.6 hrs | NR | NR | 7.11 | 3.31 – 15.25 |
|  | Hematological malignancy | NR | NR | 3.97 | 1.02 – 15.53 |
|  | Poverty stricken county residence | NR | NR | 2.35 | 1.11 – 5.00 |
| Li 2019 (b) | Need for invasive mechanical ventilation | 38.08 | 7.24 – 200.13 | 46 | 10.0 – 211.5 |
|  | PRISM ≥ 10 | 4.31 | 1.04 – 11.79 | 13.35 | 4.50 – 39.65 |
|  | Early TTP | 6.65 | 2.36 – 18.69 | 5.27 | 2.32 – 11.96 |
|  | Intensive care unit admission | NR | NR | 15.05 | 5.64 – 40.12 |
|  | Meningitis | NR | NR | 3.6 | 1.67 – 7.78 |
|  | Immunosuppression |  |  |  |  |
